# What is the best evidence for graft choice in ACL reconstruction? Protocol for a systematic review and network meta-analysis

**DOI:** 10.1101/2019.12.10.19014266

**Authors:** Junaid Habibi, Alexander Zakharia, Taylor Woolnough, Daniel Axelrod, Darren de SA

## Abstract

**Introduction:** Anterior cruciate ligament (ACL) reconstruction is one of the most commonly performed sports medicine procedures. A variety of grafts are currently used for reconstruction, including both allograft and autograft. Despite numerous meta-analyses, there exists no high-quality quantitative synthesis of all randomized controlled trial (RCT) data on graft choice.

**Objective:** To identify the optimal graft choice for ACL reconstruction by performing the first systematic review and network meta-analysis (NMA) to include both functional outcomes and complications.

**Methods:** Multiple digital databases including MEDLINE, Embase, and CENTRAL will be searched independently and in duplicate for RCTs randomizing graft choice in ACL reconstruction in skeletally mature patients. A Bayesian framework with a random-effects model will be used for NMA. Surface under the cumulative ranking curve (SUCRA) values will be used to generate a rank list for each outcome. Results will be reported as mean differences (MD) (or standardized mean difference, if necessary) or relative risk (RR) with 95% credible intervals (CI). Comparisons will be inferred to be statistically significant if the 95% CI of MD does not cross zero or if the 95% CI of relative risk does not cross one. Studies will be assessed for quality using the Cochrane risk of bias assessment tool. Quality of evidence for each network comparison will be determined as per the Grades of Recommendation, Assessment, Development and Evaluation (GRADE) approach for network meta-analyses. This NMA will be reported according to the PRISMA extension statement for network meta-analyses

**Outcomes of interest:** Functional outcomes of interest including range of motion, return to activity/sport, and IKDC, Lysholm, Tegner, ACL-QOL, and KOOS scores. Persistent laxity as measured by Lachman, Pivot-shift, side-to-side, and measured laxity (e.g. KT-1000) will also be analyzed. Complications (e.g. infection, graft failure, donor site pain), tunnel osteolysis, and failure (including but not limited to graft rupture and/or persistent laxity) will be compared between grafts.

**Relevance/Impact:** This NMA will be the first high-quality syntheses of all randomized evidence regarding graft choice in ACL reconstruction. As the first analysis to compare all major graft choices independently, it will be used to inform surgeon-patient decision making. It has the reasonable possibility of changing clinical practice.

## Introduction

Anterior cruciate ligament (ACL) reconstruction is one of the most commonly performed sports medicine procedures and the incidence of this procedure continues to increase.^1,2^ A variety of grafts have been used for reconstruction, including both allograft and autograft.^3,4^ Commonly used graft choices include bone-patellar tendon-bone autograft (BPTB), hamstrings autograft (HT), quadriceps tendon autograft (QT), and tibial tendon allograft (TT), while hybrid grafts— autograft augmented with allograft—have received recent attention.^2,3,5^ Each graft is associated with a unique functional and complication profile, often leaving surgeons and patients with the task of choosing grafts on an individualized basis.^4^ In fact, modern graft preference has remained fairly consistent with the exception of QT, which has seen a recent increase.^3,5^ Despite numerous meta-analyses and large prospective knee ligament registries, there is no clearly preferred graft, demonstrated by the frequent use of numerous different grafts.^3^ Currently, there is no high-quality quantitative synthesis of all randomized controlled trial (RCT) data on graft choice. Conventional meta-analyses are limited to comparison of two groups, necessitating either exclusion of commonly used grafts or grouping of different graft types. Network meta-analysis (NMA) generates multiple simultaneous comparisons utilizing both direct and indirect evidence.^6,7^ Accordingly, the objective of this study is to identify the optimal graft choice for ACL reconstruction by performing the first systematic review and NMA to include both functional outcomes and complications.

## Methods

This systematic review and network meta-analysis will be performed according to the Preferred Reporting Items for Systematic Reviews and Meta-Analysis (PRISMA) for network meta-analyses^8,9^ and the Cochrane Handbook for Systematic Reviews of Interventions.^10^ A PRISMA checklist for the reporting of network meta-analyses will be included as a supplementary material in the final publication.

### Literature search

A comprehensive literature search will be conducted using the following databases: Medline, Embase, CINAHL, Web of Science, Cochrane Central Register of Controlled Trials (CENTRAL), Cochrane Library, WHO International Clinical Trials Registry Platform (ICTRP), and ClinicalTrials.gov. This search will be limited to randomized controlled trials and will be conducted without language restrictions. An example search strategy for MEDLINE is provided in Table 2. The database search will be supplemented by manually screening of references from included articles and previous systematic reviews.

**Table 1.**
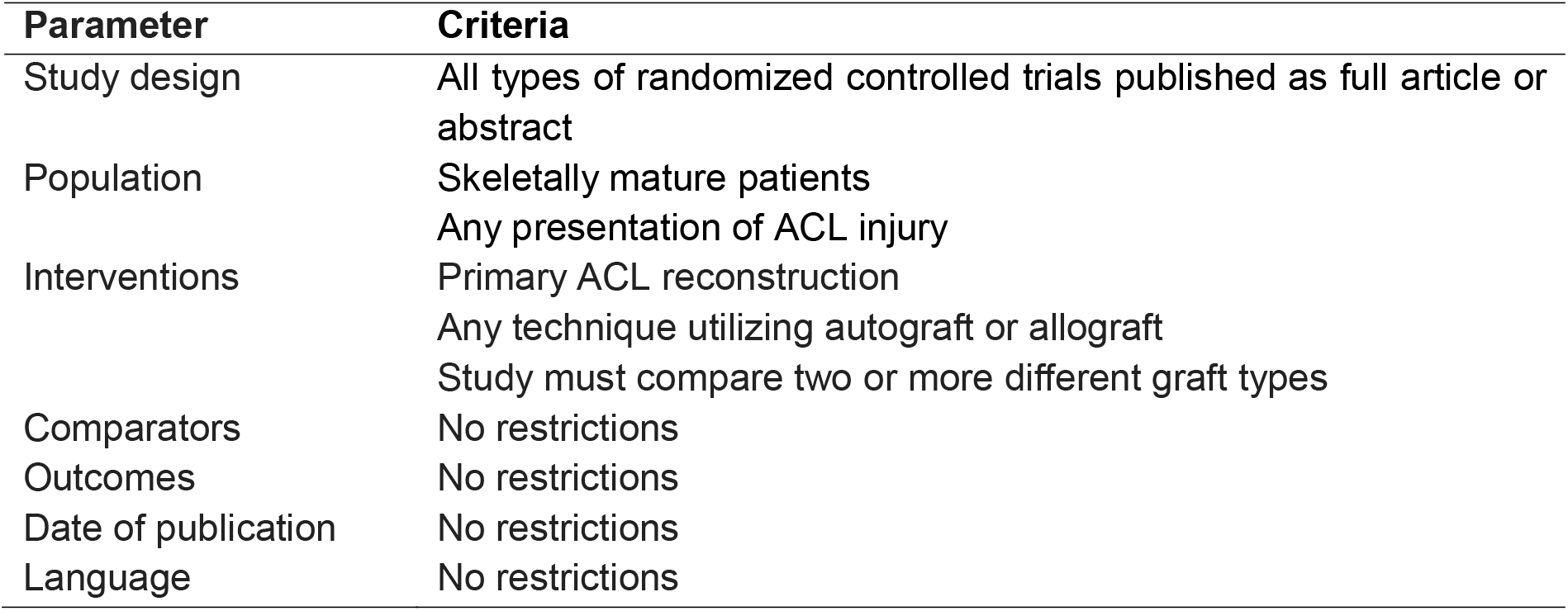
Inclusion criteria and limits

| Parameter | Criteria |
| --- | --- |
| Study design | All types of randomized controlled trials published as full article or abstract |
| Population | Skeletally mature patients<br>Any presentation of ACL injury |
| Interventions | Primary ACL reconstruction<br>Any technique utilizing autograft or allograft<br>Study must compare two or more different graft types |
| Comparators | No restrictions |
| Outcomes | No restrictions |
| Date of publication | No restrictions |
| Language | No restrictions |

**Table 2.** Search strategy for the Medline electronic database using Ovid interface for MEDLINE from 1946 to present.

| **Search Terms** |
| --- |
| 1. ACL*.mp |
| 2. Anterior cruciate ligament.mp |
| 3. Reconstruction.mp |
| 4. Graft.mp |
| 5. Single-bundle.mp |
| 6. Double-bundle.mp |
| 7. Quadriceps.mp |
| 8. Hamstring*.mp |
| 9. Patella*.mp |
| 10. 1 and 2 |
| 11. 3 or 4 or 5 or 6 or 7 or 8 or 9 |
| 12. 10 and 11 |
| 13. Limit 12 to (humans and (clinical study or clinical trial or<br>randomized controlled trial)) |

**Table 3.** Cochrane risk of bias evaluation criteria

| Domain | Risk of Bias | Justification |
| --- | --- | --- |
| Random sequence generation (selection bias) | Low | Randomisation and its method is determined to be unpredictable. |
|  | High | No randomisation reported or states a predictable randomisation method. |
|  | Unclear | The study does not report the randomisation method, but mentions explicitly that randomisation occurred or the study describes allocation concealment but does not discuss the method of randomisation. |
| Allocation concealment (selection bias) | Low | Method of randomisation reported and determined to be difficult to circumvent. |
|  | High | Method of randomisation is reported to be relatively easy to circumvent. |
|  | Unclear | The study does not report the method used to maintain the integrity of randomisation or the study describes randomisation method without discussing how allocation concealment was maintained. |
| Blinding of participants and personnel (performance bias) | Low | The study discusses that blinding occurred either explicitly or implicitly. Blinding of surgeon is not possible. Patient blinding will be determined by reporting that patients were blinded. |
|  | High | The study does not discuss blinding either implicitly or explicitly. |
|  | Unclear | Only low and high risk of bias will be used for this domain. |
| Blinding of outcome assessment (detection bias) | Low | The study discusses blinding of outcome assessor either explicitly or implicitly. |
|  | High | The study does not mention outcome assessor blinding implicitly or explicitly. |
|  | Unclear | The study discusses that blinding occurred but does not mention blinding of the outcome assessor. |
| Incomplete outcome data (attrition bias) | Low | <20% loss to follow-up; or loss to follow-up not related to intervention; or use of “intention-to-treat” analysis. |
|  | High | Loss to follow-up ≥20% and no mention of loss to follow-up reason; or loss to follow-up related to intervention; or use of “as-treated” analysis. |
|  | Unclear | No mention of follow-up. |
| Selective reporting bias (reporting bias) | Low | Prespecified outcomes in methods were reported in results; prespecified outcomes in protocol were reported in results. |
|  | High | Prespecified outcomes in methods were not reported in results; prespecified outcomes in protocol were not reported in results. |
|  | Unclear | Outcomes unclear. |
| Other sources of bias (industry sponsorship) | Low | No industry sponsorship. |
|  | High | Industry sponsorship or other concerns. |
|  | Unclear | Unclear disclosure of industry sponsorship. |

From: Higgins JPT, Altman DG, Gøtzsche PC, et al. The Cochrane Collaboration's tool for assessing risk of bias in randomised trials. *BMJ*. 2011;343:d5928.

**Table 4.**
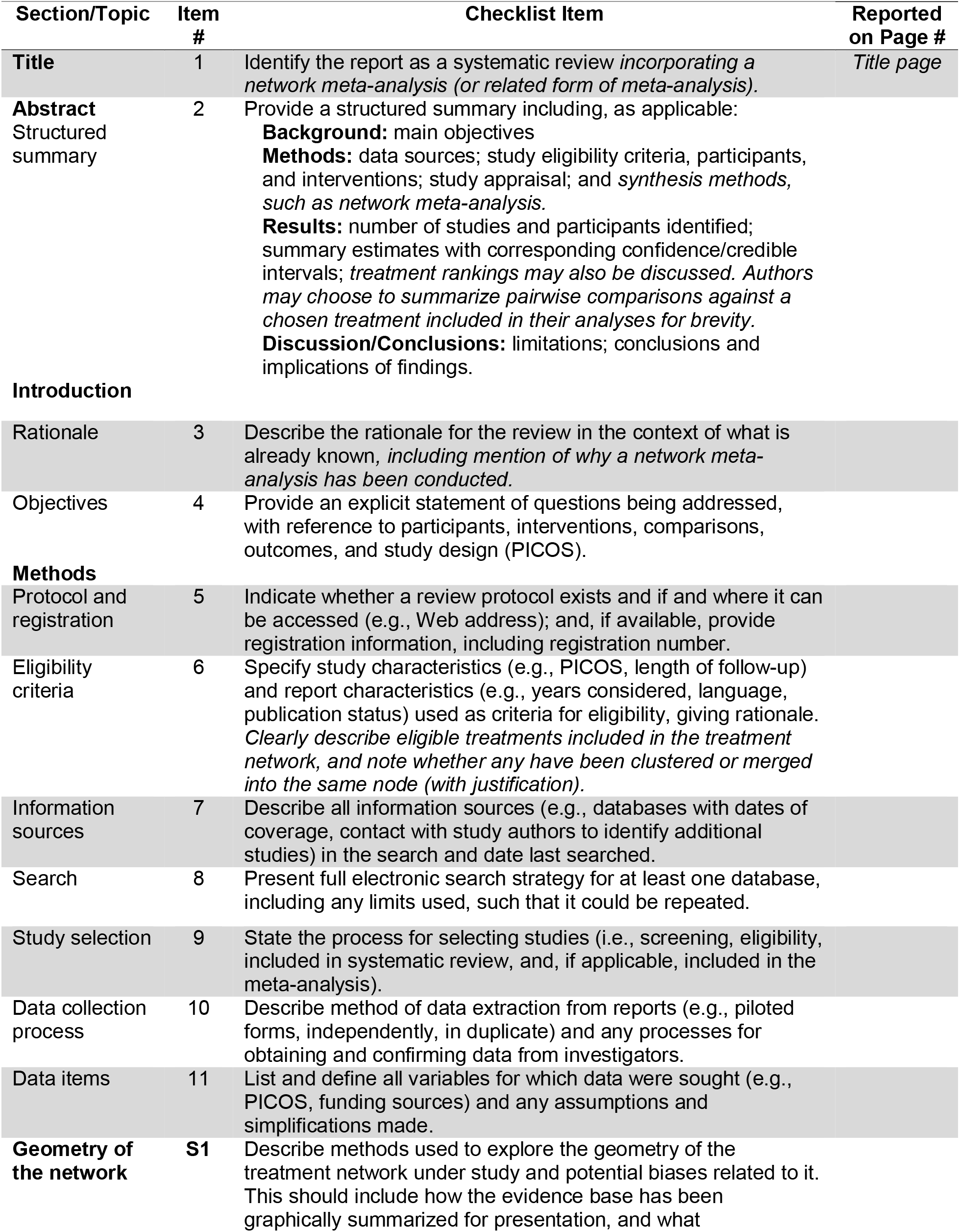
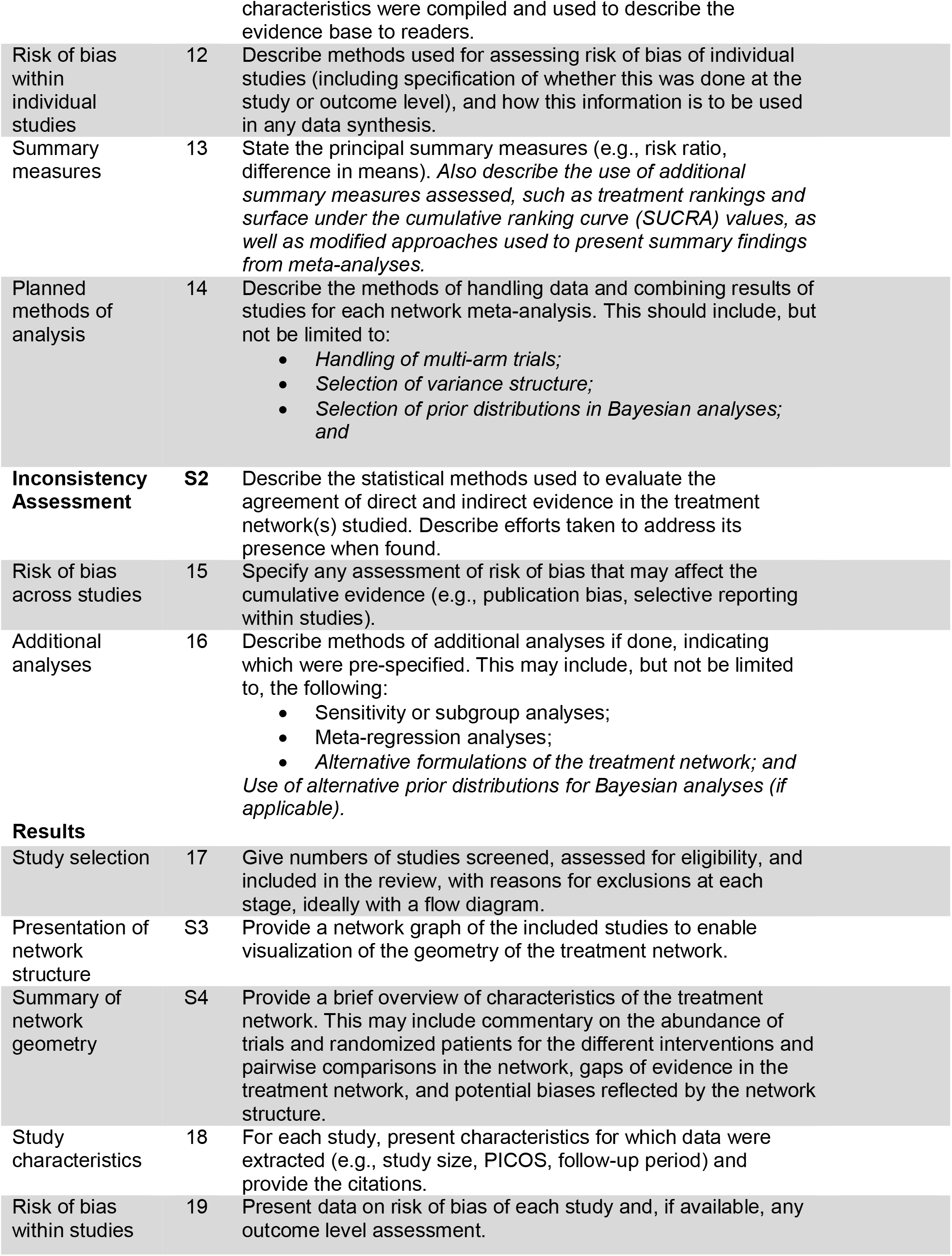
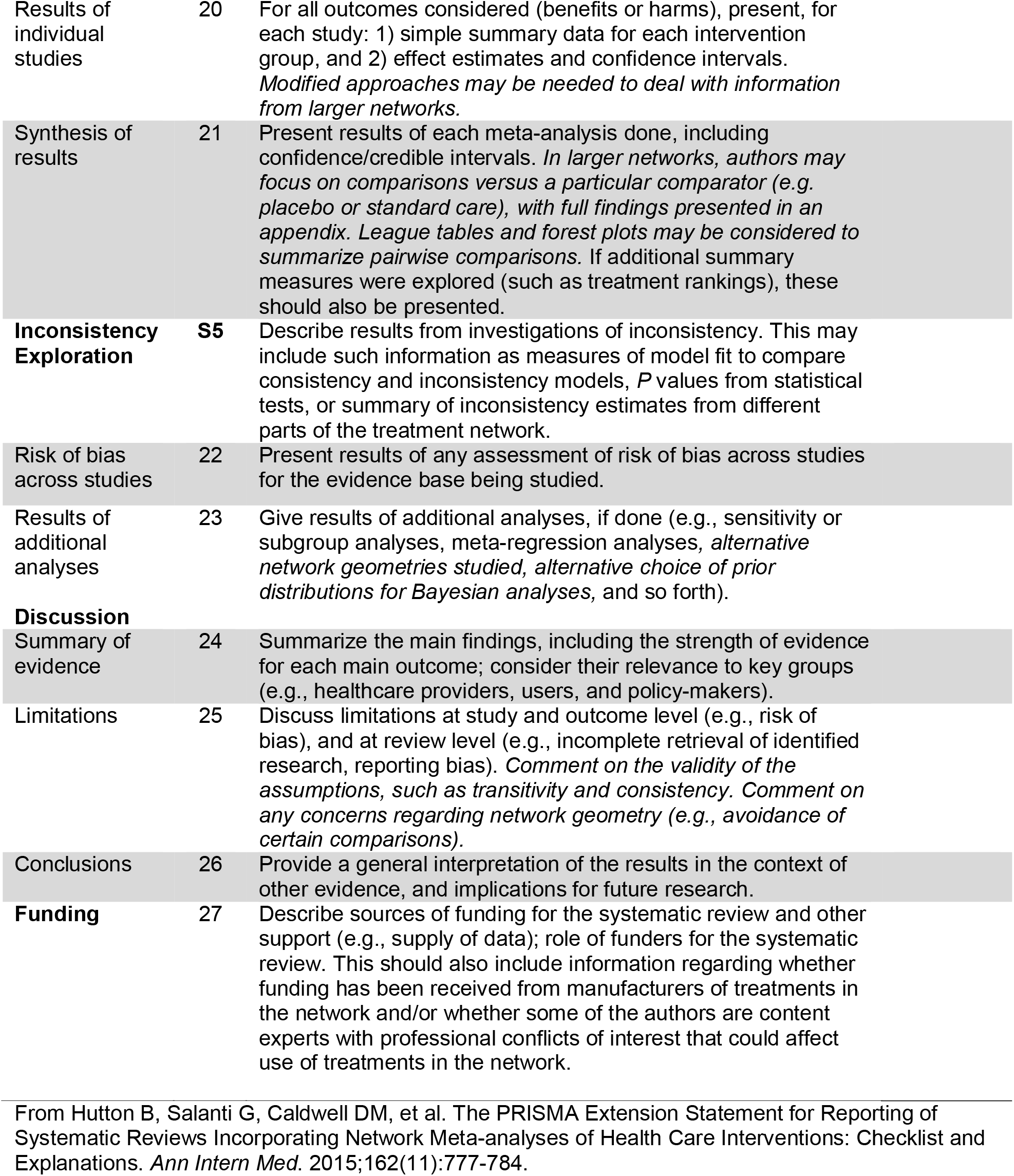
PRISMA Systematic Review and Network Meta-Analysis Reporting Checklist

| Section/Topic | Item # | Checklist Item | Reported on Page # |
| --- | --- | --- | --- |
| **Title** | 1 | Identify the report as a systematic review *incorporating a network meta-analysis (or related form of meta-analysis)*. | *Title page* |
| **Abstract**<br>Structured summary | 2 | Provide a structured summary including, as applicable:<br>**Background:** main objectives<br>**Methods:** data sources; study eligibility criteria, participants, and interventions; study appraisal; and *synthesis methods, such as network meta-analysis*.<br>**Results:** number of studies and participants identified; summary estimates with corresponding confidence/credible intervals; *treatment rankings may also be discussed*. *Authors may choose to summarize pairwise comparisons against a chosen treatment included in their analyses for brevity*.<br>**Discussion/Conclusions:** limitations; conclusions and implications of findings. |  |
| **Introduction** |  |  |  |
| Rationale | 3 | Describe the rationale for the review in the context of what is already known, *including mention of why a network meta-analysis has been conducted*. |  |
| Objectives | 4 | Provide an explicit statement of questions being addressed, with reference to participants, interventions, comparisons, outcomes, and study design (PICOS). |  |
| **Methods** |  |  |  |
| Protocol and registration | 5 | Indicate whether a review protocol exists and if and where it can be accessed (e.g., Web address); and, if available, provide registration information, including registration number. |  |
| Eligibility criteria | 6 | Specify study characteristics (e.g., PICOS, length of follow-up) and report characteristics (e.g., years considered, language, publication status) used as criteria for eligibility, giving rationale. *Clearly describe eligible treatments included in the treatment network, and note whether any have been clustered or merged into the same node (with justification)*. |  |
| Information sources | 7 | Describe all information sources (e.g., databases with dates of coverage, contact with study authors to identify additional studies) in the search and date last searched. |  |
| Search | 8 | Present full electronic search strategy for at least one database, including any limits used, such that it could be repeated. |  |
| Study selection | 9 | State the process for selecting studies (i.e., screening, eligibility, included in systematic review, and, if applicable, included in the meta-analysis). |  |
| Data collection process | 10 | Describe method of data extraction from reports (e.g., piloted forms, independently, in duplicate) and any processes for obtaining and confirming data from investigators. |  |
| Data items | 11 | List and define all variables for which data were sought (e.g., PICOS, funding sources) and any assumptions and simplifications made. |  |
| **Geometry of the network** | S1 | Describe methods used to explore the geometry of the treatment network under study and potential biases related to it. This should include how the evidence base has been graphically summarized for presentation, and what |  |

|  |  |  |
| --- | --- | --- |
|  |  | characteristics were compiled and used to describe the evidence base to readers. |
| Risk of bias within individual studies | 12 | Describe methods used for assessing risk of bias of individual studies (including specification of whether this was done at the study or outcome level), and how this information is to be used in any data synthesis. |
| Summary measures | 13 | State the principal summary measures (e.g., risk ratio, difference in means). *Also describe the use of additional summary measures assessed, such as treatment rankings and surface under the cumulative ranking curve (SUCRA) values, as well as modified approaches used to present summary findings from meta-analyses.* |
| Planned methods of analysis | 14 | Describe the methods of handling data and combining results of studies for each network meta-analysis. This should include, but not be limited to:<br>• *Handling of multi-arm trials;*<br>• *Selection of variance structure;*<br>• *Selection of prior distributions in Bayesian analyses; and* |
| **Inconsistency Assessment** | **S2** | Describe the statistical methods used to evaluate the agreement of direct and indirect evidence in the treatment network(s) studied. Describe efforts taken to address its presence when found. |
| Risk of bias across studies | 15 | Specify any assessment of risk of bias that may affect the cumulative evidence (e.g., publication bias, selective reporting within studies). |
| Additional analyses | 16 | Describe methods of additional analyses if done, indicating which were pre-specified. This may include, but not be limited to, the following:<br>• Sensitivity or subgroup analyses;<br>• Meta-regression analyses;<br>• *Alternative formulations of the treatment network; and*<br>*Use of alternative prior distributions for Bayesian analyses (if applicable).* |
| **Results** |  |  |
| Study selection | 17 | Give numbers of studies screened, assessed for eligibility, and included in the review, with reasons for exclusions at each stage, ideally with a flow diagram. |
| Presentation of network structure | S3 | Provide a network graph of the included studies to enable visualization of the geometry of the treatment network. |
| Summary of network geometry | S4 | Provide a brief overview of characteristics of the treatment network. This may include commentary on the abundance of trials and randomized patients for the different interventions and pairwise comparisons in the network, gaps of evidence in the treatment network, and potential biases reflected by the network structure. |
| Study characteristics | 18 | For each study, present characteristics for which data were extracted (e.g., study size, PICOS, follow-up period) and provide the citations. |
| Risk of bias within studies | 19 | Present data on risk of bias of each study and, if available, any outcome level assessment. |
| Results of individual studies | 20 | For all outcomes considered (benefits or harms), present, for each study: 1) simple summary data for each intervention group, and 2) effect estimates and confidence intervals. *Modified approaches may be needed to deal with information from larger networks.* |
| Synthesis of results | 21 | Present results of each meta-analysis done, including confidence/credible intervals. *In larger networks, authors may focus on comparisons versus a particular comparator (e.g. placebo or standard care), with full findings presented in an appendix. League tables and forest plots may be considered to summarize pairwise comparisons.* If additional summary measures were explored (such as treatment rankings), these should also be presented. |
| **Inconsistency Exploration** | **S5** | Describe results from investigations of inconsistency. This may include such information as measures of model fit to compare consistency and inconsistency models, *P* values from statistical tests, or summary of inconsistency estimates from different parts of the treatment network. |
| Risk of bias across studies | 22 | Present results of any assessment of risk of bias across studies for the evidence base being studied. |
| Results of additional analyses | 23 | Give results of additional analyses, if done (e.g., sensitivity or subgroup analyses, meta-regression analyses, *alternative network geometries studied, alternative choice of prior distributions for Bayesian analyses, and so forth*). |
| **Discussion** |  |  |
| Summary of evidence | 24 | Summarize the main findings, including the strength of evidence for each main outcome; consider their relevance to key groups (e.g., healthcare providers, users, and policy-makers). |
| Limitations | 25 | Discuss limitations at study and outcome level (e.g., risk of bias), and at review level (e.g., incomplete retrieval of identified research, reporting bias). *Comment on the validity of the assumptions, such as transitivity and consistency. Comment on any concerns regarding network geometry (e.g., avoidance of certain comparisons).* |
| Conclusions | 26 | Provide a general interpretation of the results in the context of other evidence, and implications for future research. |
| **Funding** | 27 | Describe sources of funding for the systematic review and other support (e.g., supply of data); role of funders for the systematic review. This should also include information regarding whether funding has been received from manufacturers of treatments in the network and/or whether some of the authors are content experts with professional conflicts of interest that could affect use of treatments in the network. |

From Hutton B, Salanti G, Caldwell DM, et al. The PRISMA Extension Statement for Reporting of Systematic Reviews Incorporating Network Meta-analyses of Health Care Interventions: Checklist and Explanations. *Ann Intern Med.* 2015;162(11):777-784.

### Screening

Articles will be screened by two investigators, independently and in duplicate. Disagreements between reviewers will be resolved by consensus, and if necessary, consultation with a senior reviewer. For studies published more than once, only the article with the most complete data will be included. For articles published in a non-English language, a medical translator fluent in the language of the study will translate and assist with article screening if possible.

Exclusion criteria will include trials randomizing interventions other than graft choice (e.g. rehabilitation protocol, graft fixation method), observational studies, case series, case reports, biomechanical cadaver studies, review articles, letters, basic science studies, correspondences or comments. Trials examining revision reconstruction or those including skeletally immature patients will be excluded.

### Data Extraction

Articles produced from the literature search will be downloaded as complete reference files. Two reviewers will independently extract data in parallel from all included articles into a piloted, standard extraction document (Microsoft Excel 16.2, Redmond, United States) designed *a priori*. Article authors will be contacted in instances where additional information or clarity is required. The following study details will be extracted: year of publication, study design, recruitment period, country of recruitment, trial funder, role of trial funder, number of patients, number of knees, patient age, sex, and activity level, graft fixation method, concurrent procedures at the time of ACL reconstruction, post-operative protocol, and follow-up duration.

### Outcomes of Interest

Outcomes of interest include range of motion, return to activity/sport, subjective and objective International Knee Documentation Committee (IKDC)^11,12^, Lysholm^13,14^, ACL-Quality of Life (ACL-QOL)^2^, Knee Injury and Osteoarthritis Outcome (KOOS)^15^, and Tegner scores^14,16^, results of Lachman and Pivot-shift tests, side-to-side, measured laxity (e.g. KT-1000), complications (e.g. infection, graft failure, donor site pain), tunnel osteolysis, and failure (including but not limited to graft rupture and/or persistent laxity).

If range of motion is reported only as a percentage of the uninjured knee, a degree measurement will be calculated using published normal values.^17,18^ Means and standard deviations (SDs) will be collected; medians will be used in lieu of means if mean values are not reported.^19^ If a 95% confidence interval was reported as the measure of variability, standard deviation will be approximated.^20^ When no measure of variance is reported, standard deviation will be imputed using a p value (if reported exactly) or a weighted average of variances observed in other included studies.^20,21^

### Statistical Analysis

Statistical analysis was performed using R 3.4.2 (Open Access Online) with BUGSnet (Lighthouse Outcomes, Toronto, Canada) and CINeMA.^22^ Heterogeneity between studies will be calculated using the I^2^ statistic; if heterogeneity is high, a Bayesian framework with a random-effects model and non-informative priors will be used. A graphical framework of all trials comparing different interventions will be created for each outcome. Ranking diagrams and forest plots will also be created for each outcome. Furthermore, surface under the cumulative ranking curve (SUCRA) values will be reported for each study. The SURCRA score represents the likelihood that a given treatment will rank first in a specific category; a score closer to one indicates that treatment is more likely to represent the best treatment. Treatment rank orders will be generated and presented using SUCRA values. Incoherence—inconsistency between direct and indirect evidence—will be assessed globally using the design-by-treatment interaction test and individually using the Separating Indirect from Direct Evidence (SIDE) and node-splitting methods.^23,24^ If appropriate, a nested analysis will be conducted; the first level of analysis will compare autograft to allograft while the second level of analysis will look at each graft independently.

Results of the network for functional outcomes will be reported as mean differences (MD) (or standardized mean difference, if necessary) with 95% credible intervals (CI). Complications will be presented using relative risk (RR), 95% credible intervals, and number needed to treat, as appropriate. Credible intervals are derived using the posterior distribution of the outcome in question and can be thought of as the Bayesian equivalent of confidence intervals. Comparisons will be inferred as statistically significant if the 95% CI of MD does not cross zero or if the 95% CI of relative risk does not cross one.

### Quality Assessment

The quality of each included study will be evaluated in duplicate using the Cochrane Risk of Bias tool, while the Cochrane CINeMA tool, designed around the GRADE framework, will be used for risk of bias assessment specific to outcomes from network meta-analyses.^22,25^ Included RCTs will be assessed for quality by two independent reviewers using the Cochrane risk of bias assessment tool.^26^ Disagreement will be resolved through consultation with a third reviewer. Overall quality of evidence for each network comparison will be determined and ranked based on within-study bias, reporting bias, indirectness, imprecision, heterogeneity, and incoherence, as per the Grades of Recommendation, Assessment, Development and Evaluation (GRADE) approach for network meta-analyses.^25,27^

## Data Availability

Not applicable.

## ETHICS AND DISSEMINATION

Approval from a research ethics board is not required as this study will synthesize data from conducted studies. Results from this study are expected to comprehensively summarize and inform clinical practice. The results from this review will be submitted to a peer-reviewed journal for publication and will be presented at conferences.

## AUTHORS’ CONTRIBUTIONS

TW, DA, and DdS conceived and designed the protocol. AZ and JH designed the search strategy and piloted it across all relevant databases. TW, DA, and DdS developed the review protocol, selection criteria, and risk of bias assessment. TW and DA designed the data management and synthesis methodology. AZ and JH performed data abstraction. All authors critically reviewed and collaborated in the discussion of the intellectual content of the protocol. All authors provided final approval of the protocol.

